# The potential impact of removing a ban on electronic nicotine delivery systems using the Mexico smoking and vaping model (SAVM)

**DOI:** 10.1101/2024.04.28.24306511

**Authors:** Luz María Sánchez-Romero, Yameng Li, Luis Zavala-Arciniega, Katia Gallegos-Carrillo, James F. Thrasher, Rafael Meza, David T. Levy

## Abstract

**Objective:** To develop the Mexico Smoking and Vaping Model (Mexico SAVM) to estimate cigarette and electronic nicotine delivery systems (ENDS) prevalence and the public health impact of legalizing ENDS use.

**Methods:** SAVM, a cohort-based discrete-time simulation model, compares two scenarios. The *ENDS-Restricted Scenario* estimates smoking prevalence and associated mortality outcomes under the current policy of an ENDS ban, using Mexico-specific population projections, death rates, life expectancy, and smoking and e-cigarette prevalence. *The ENDS-Unrestricted Scenario* projects smoking and vaping prevalence under a hypothetical scenario where ENDS use is allowed. The impact of legalizing ENDS use is estimated as the difference in smoking- and vaping-attributable deaths (SVADs) and life-years lost (LYLs) between the ENDS-Restricted and Unrestricted scenarios.

**Results:** Compared to a national ENDS ban, The Mexico SAVM projects that legalizing ENDS use could decrease smoking prevalence by 40.1% in males and 30.9% in females by 2049 compared to continuing the national ENDS ban. This reduction in prevalence would save 2.9 (2.5 males and 0.4 females) million life-years and avert almost 106 (91.0 males and 15.5 females) thousand deaths between 2025 and 2049. Public health gains decline by 43% to 59,748 SVADs averted when the switching rate is reduced by half and by 24.3% (92,806 SVADs averted) with a 25% ENDS risk level from that of cigarettes but increased by 24.3% (121,375 SVADs averted) with the 5% ENDS risk.

**Conclusions:** Mexico SAVM suggests that greater access to ENDS and a more permissive ENDS regulation, simultaneous with strong cigarette policies, would reduce smoking prevalence and decrease smoking-related mortality. The unanticipated effects of an ENDS ban merit closer scrutiny, with further consideration of how specific ENDS restrictions may maximize public health benefits.

## Introduction

Patterns of tobacco use worldwide have changed considerably over time (1–3). In the last decade, many high-income countries (HICs) have seen rapid growth in the use of electronic nicotine delivery systems (ENDS, aka e-cigarettes), especially among younger people. However, ENDS use is much less prevalent in low and middle-income countries (LMICs) (4–13). Compared to HICs, LMICs tend to have relatively low rates of ENDS use among people who have never smoked cigarettes but relatively high rates of “dual use” with cigarettes (4–10).

Currently, 34 countries – representing 17% of LMICs and 13% of HICs – have banned ENDS (14, 15), with some evidence for their effectiveness (16–19). However, enforcement of these bans in LMICs is often weak, and implementation is frequently challenged by the relatively large size of informal economies in those countries. In contrast, some HICs have adopted permissive approaches to regulating ENDS. This approach is driven by the recognition of ENDS’ potential for harm reduction, especially among adults who struggle to quit cigarette smoking (20). The potential of ENDS for harm reduction is likely to be greater when implemented alongside strong regulations to discourage cigarette use, which, in general, tend to be stronger in HICs than LMICs (14). A simulation model for Australia, one of the few HICs with strict ENDS regulations and strong cigarette-oriented policies, reported that a permissive ENDS policy could reduce smoking and ENDS associated deaths by 7.7% compared to the current restrictive ENDS regulation (21). A modeling study for New Zealand projected that removing their ENDS ban could gain 236,000 quality-adjusted life years (QALYs) and save US$2.5 billion (22). Simulating the effects of alternative regulatory approaches in LMICs that ban ENDS has yet to be done but could offer valuable insights into the need to tailor regulations to specific country contexts.

This paper aims to develop a population simulation model that considers the impact of shifting from stricter to more permissive ENDS policies in LMICs. Currently, seven countries in Latin America, of which six are LMICs, have banned ENDS (15). We focus on Mexico as a case study for LMICs removal of a current ENDS ban because of its large population, implementation of a range of WHO-recommended tobacco control policies (e.g., taxes 69% of the final price (23); smoke-free areas (24, 25); pictorial warnings on packs (26, 27); marketing bans), and relatively stable smoking prevalence over the period since their implementation (28). The federal-level Tobacco Control Act of 2008 included a de facto ban on the importation, distribution, marketing and sales of novel nicotine products, which regulators interpreted as encompassing ENDS; courts affirmed the ban in 2021 and 2022 (29). Current e-cigarette use (exclusive and dual) among 15- to 65-year-old Mexicans has increased from 0.7% in 2015 to 1.5% in 2018 and has remained near that level through 2022 (30). However, from 2015-2022, male and female exclusive e-cigarette use increased (males=0.40% to 1.9%; females=0.14%- 0.83%) while dual use of ENDS with cigarettes also increased (males=0.77% to 1.46%; females=0.12% to 0.74%) (31). Meanwhile, current e-cigarette use among adolescents ages 15 to 19 years increased from 2.1% in 2015 to 2.6% in 2022 (30).

We developed the Mexico Smoking and Vaping Model (SAVM) based on the structure of the “Australia SAVM” (21). The Mexico SAVM projects cigarette and ENDS prevalence with and without an ENDS ban and calculates the public health impact of legalizing ENDS use. The model offers preliminary evidence for decision-makers about the potential effects of lifting Mexico’s ENDS ban on product use rates and tobacco-related mortality.

## Materials and methods

The Mexico SAVM is a cohort-based discrete-time simulation model that simulates cigarette smoking and vaping for the Mexican population and estimates the public health impact of legalizing ENDS use (vaping) by simulating and comparing two scenarios. First, the *ENDS-Restricted Scenario* (baseline) estimates smoking prevalence and associated mortality outcomes when vaping is restricted and accounts for ENDS use under those restrictions (due to limited enforcement of tobacco control regulations) (32, 33). Second, *the ENDS-Unrestricted Scenario* incorporates permissive ENDS access into the cohort trajectories to project smoking and vaping prevalence in a hypothetical scenario where ENDS use is allowed, or access is less restricted, similarly to their status in the US until 2021. The public health impact is then derived by comparing smoking- and vaping-attributable mortality and averted life-years lost outcomes in the ENDS-Unrestricted Scenario (less stringent ENDS regulation) to those in the ENDS-Restricted Scenario.

The Mexico SAVM runs from 2009 to 2049 and simulates individuals ages 15 to 65. We focus our analysis on individuals up to 65 years old to capture a demographic where smoking behaviors are likely most influenced by using non-cigarette nicotine products: the consumption of ENDS among Mexican adults aged 65 and above is negligible (0.1% current ENDS users) (34). The 2009 baseline was chosen as the initial year based on the availability of a nationally representative survey to initialize the model and relatively stable tobacco control policy levels in the years before and after 2009.(33) We decided on a short modeling period because the latest available nationally representative smoking data are from 2020-2022. The lockdowns during the COVID-19 pandemic (particularly in 2020 and 2021) impacted the population’s health-related behaviors. Thus, data from that period or immediately afterward may not represent reliable smoking and vaping trends (35–37). Table 1 lists the demographic data sources and parameter values used as inputs in the Mexico SAVM.

**Table 1:** Data sources and parameter values used in the Mexico SAVM.

| Input parameters | Description | Data source or estimate |
| --- | --- | --- |
| Population | Population by age, gender, and year (2009-2049) | The Mexico National Council of Population (CONAPO). |
| Mortality rates for overall population | Overall mortality rates by age, and gender in 2009; separated by the US overall and smoking status-specific death rate; extended using US smoking status trend (2009-2049). | The Mexico National Council of Population (CONAPO). |
| Expected life years for overall population | Overall life expectancy by age, gender, scaled up by the US never smoking and overall life expectancy ratio; extended using US trend (2009-2049). | Global Health Observatory. Mexico-Life expectancy and healthy life expectancy |
| Smoking prevalence | Prevalence of never, current, and former smokers by age group and gender for initial year | 2009 Global Adult Tobacco Survey (GATS), Mexico |
| Smoking initiation and cessation rate for the ENDS-Restricted Scenario | Percentage of never smokers who become smokers and the percentage of smokers who permanently quit every year. | Patterns of Birth Cohort-Specific Smoking Histories in Mexico |
| ENDS users from Never Smokers (ENDS-PN) | Proportion of ENDS users among never smokers | Global Adult Tobacco Survey (GATS) in 2015, the National Survey of Drugs, Alcohol, and Tobacco Consumption (ENCODAT) in 2016 and the National Health and Nutrition Survey (ENSANUT) in 2018, 2020, 2021 and 2022 |
| ENDS users from former smokers (ENDS-PF) | Proportion of ENDS users among former smokers |  |
| Smoking initiation adjustor for the ENDS-Restricted Scenario | Ratio of smoking initiation rate in the Restricted Scenario | Male (female):<br>15-24 years: 1.52 (1.53)<br>25-40 years: 1.78 (1.56) |
| Smoking cessation adjustor for the ENDS-Restricted Scenario | Ratio of smoking cessation rate in the Restricted Scenario | Male (female):<br>25-39 years: 0.50 (0.50)<br>40+ years: 0.48(0.00) |
| ENDS relative risk multiplier | Excess risk of ENDS use measured relative to excess smoking risks (mortality rate | ENDS excess mortality risks set at 15% (5% and 25%)<sup>†</sup> |
|  | of current smokers – mortality rate of never smokers) |  |
| ENDS Switching rate | Rate of switching from smoking cigarettes to exclusive ENDS use | Ranges from 0.3%-2.0%, estimated by age group and gender using the 50% of the estimated switching rate from a prospective analysis from PATH data wave 1 (2013) to wave 4 (2017) |
| Smoking initiation multiplier in the ENDS-Unrestricted Scenario | Ratio of smoking initiation rate in the Unrestricted Scenario to smoking initiation rate in the Restricted Scenario | 75% of the Restricted Scenario smoking initiation rate, based on recent studies.(59-61) |
| ENDS initiation multiplier in the ENDS-Unrestricted Scenario | Ratio of ENDS initiation rate in the Unrestricted Scenario to smoking initiation rate in the Restricted Scenario | 50% of the Restricted Scenario smoking initiation rate, based on recent studies.(98-101) |
| Smoking cessation multiplier in the ENDS-Unrestricted Scenario | Ratio of smoking cessation rate in the Unrestricted Scenario to smoking cessation rate in the Restricted Scenario | 100% of the ENDS-Restricted Scenario smoking cessation rate |
| ENDS cessation multiplier in the ENDS-Unrestricted Scenario | Ratio of ENDS cessation rate in the Unrestricted Scenario to smoking cessation rate in the Restricted Scenario | 100% of the ENDS-Restricted Scenario smoking cessation rate |

ENDS=electronic nicotine delivery systems, *ENDS-Restricted Scenario*=values in the presence of an ENDS ban. *ENDS-Unrestricted Scenario*=values in the absence of an ENDS ban. PATH= The Population Assessment of Tobacco and Health Study, US. <sup>\*</sup>Suggested change in ENDS relative risk multiplier for sensitivity analysis.

### The ENDS-Restricted Scenario

The ENDS-Restricted Scenario estimates the prevalence and mortality of never, current (daily and nondaily), and former smokers, as well as never smokers using ENDS (N-ENDS) and former smokers using ENDS (FS-ENDS) over time in the present Mexican scenario of an ENDS ban, a nationally implemented sales ban with low enforcement (Fig 1).

**Fig 1.**
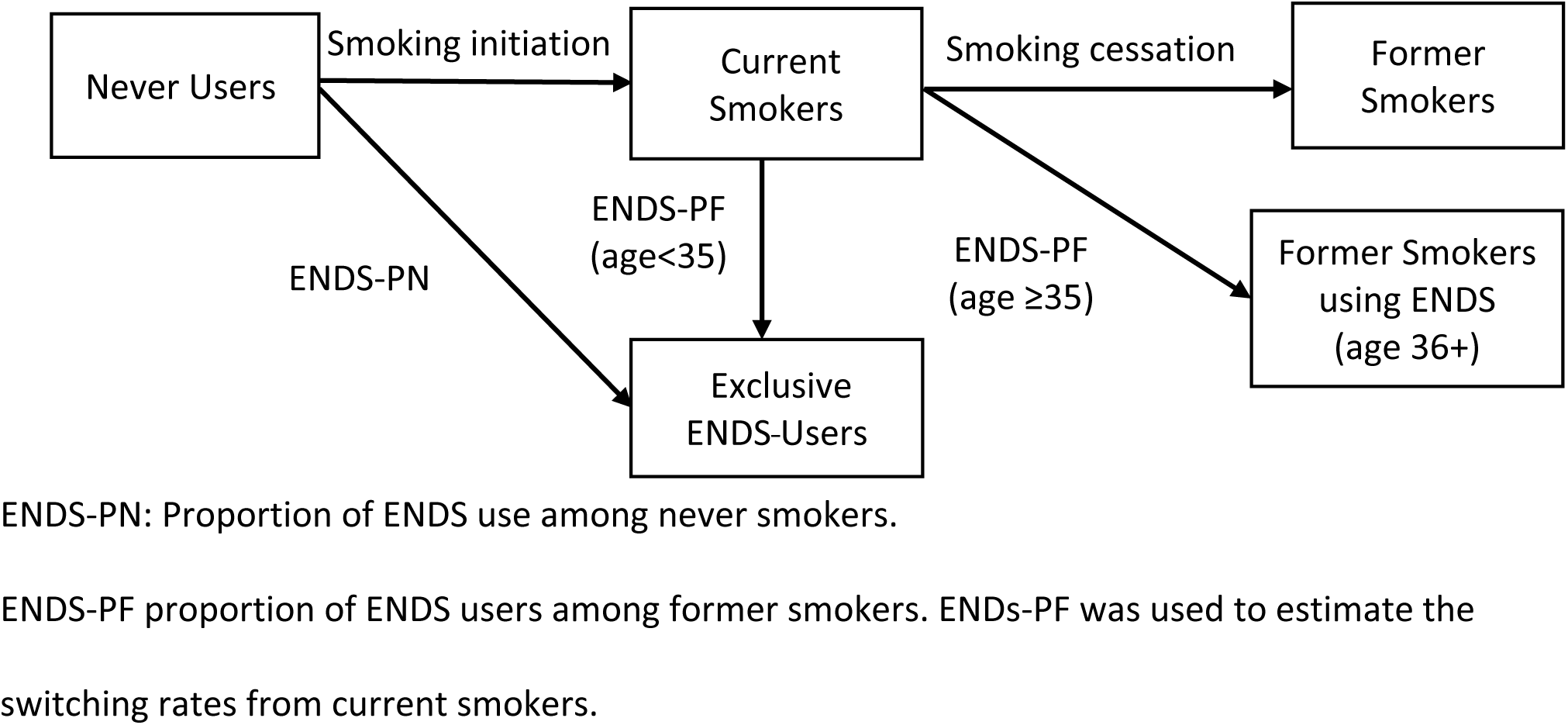
Transitions between Smoking and ENDS Use in the ENDS-Restricted Scenario

SAVM projects the population by simulating separate cohorts for males and females by individual age. Births and overall mortality are based on Mexico’s single-age yearly population projections for 2009-2049 from the Mexico National Council of Population (CONAPO) (38, 39). Gender-specific life expectancy by five-year age groups (ages 1 to 85+) for 2010, 2015 and 2019 from the World Health Organization-Global Health Observatory (40). Since Mexico’s life expectancy was only available for certain years and SAVM requires life-expectancy data for the complete modeling period, we applied the 2010 life expectancy as the same life expectancy in 2009-2014, the 2015 as the life expectancy in 2015-2018, and the 2019 rates as extrapolated to 2049. Mexico’s overall life expectancy was then transformed into never smokers’ life expectancy for each modeling year by multiplying this data by the ratio of the 2016 US never smoking and US overall life expectancy (41), assuming that each age-gender ratio is constant over the modeling period. US Life expectancy of never smokers increases over time in relative terms by 7% (5%) from 2009-2049 for males (females) at age 20, 14% (10%) for age 40, and 22% (18%) for age 60.

Mexico-SAVM smoking prevalence estimates are based on Mexico’s prevalence by smoking status in the initial year (2009) and Mexico-specific smoking initiation and cessation rates. Mexico SAVM uses gender-specific weighted smoking status prevalence (never, current, and former) by five-year age groups (ages 15-65) from the nationally representative 2009 Global Adult Tobacco Survey (GATS)-Mexico (42). GATS defines current smokers as those individuals who answered “daily” and “less than daily” to the question “Do you currently smoke tobacco on a daily basis, less than daily, or not at all?”. Former smokers were those who responded, “Not at all” to “Do you currently smoke tobacco on a daily basis, less than daily, or not at all?” and “daily or less than daily” to “In the past have you smoked tobacco on a daily basis, less than daily or not at all?” Never users are those who never smoked tobacco products or ENDS in their lifetime. GATS considers smokers as those who smoke any combustible tobacco (cigars, cigarettes, and pipes) either daily or occasionally. However, in Mexico, among all the people who smoke tobacco, about 98-99% smoke cigarettes (43). Therefore, Mexico SAVM assumes smoking corresponds to cigarette use. Further, SAVM does not distinguish between dual use (smoking and ENDS) and exclusive smokers due to the relatively unstable transitions of dual use behaviors and limited information. Most relapse by former smokers occurs within one year after quitting. Relapse among those who quit for ≥2 years is less than 30% (44–47). Permanent cessation is measured by quitting for at least two years to reflect cessation net of relapse rather than distinguishing former smoking by years quit.

To fit the model by single-age smoking status prevalence in the initial year, we transformed the 2009 GATS five-year age groups (ages 15 to 65 years) prevalence into individual age prevalence by assigning the prevalence by age group to the mid-age of each age group and then linear interpolating between those mid-ages (SAVM User Guide, sec 3.2.4). For example, we apply the same prevalence for ages groups 15 to 19 and 20 to 24 as the prevalence at ages 17 and 22 (the midpoints in the 15-19 and 20-24 age groups from GATS) and estimate the prevalence at ages 18-21 by assuming the prevalence at age 17 will evenly and gradually increase to the prevalence at age 22. Current and former smoking prevalence is set to 0% before age 9 and age 15, respectively. After age 62 (the mid-age of the last age group 60-65), we assume the prevalence for older ages is the same as the prevalence at age 62.

The Mexico initiation and cessation rates (by single age 0-65), gender and single year 2009-2049) were estimated using age-period cohort models and national survey data following the same approach as previous analyses for the US (48, 49). For initiation, we used data from Mexico’s national health surveys [National Survey on Drug Use and Health (ENA), National Health and Nutrition Survey (ENSANUT) and GATS) covering 1998-2016 and reporting data on age at initiation from ever smokers. For cessation, we used data from national surveys covering the period 1998-2016 and reporting on age at cessation for former smokers (ENA and GATS). Based on these estimates, Mexico SAVM applies initiation rates from ages 6 to 39 years and cessation rates from ages 11 to 65.

To reflect current Mexico ENDS use despite the ban (31), Mexico SAVM incorporated ENDS use under the ENDS Restricted Scenario starting in 2015 and through the modeling period. In this scenario, Mexico SAVM assumes that exclusive ENDS users come from never smokers using ENDS (N-ENDS) and former smokers who use ENDS (FS-ENDS). SAVM considers N-ENDS users as those who initiate exclusive ENDS daily or less than daily in the absence of smoking for all ages and those who quit cigarette use before age 35 and switch to exclusive ENDS use. FS-ENDS are those who quit cigarette use after age 35 and switch to exclusive ENDS use. To estimate N-ENDS and FS-ENDS, we used the proportion of ENDS use among never smokers (ENDS-PN) and the proportion of ENDS users among former smokers (ENDS-PF) from Mexican national datasets. The ENDS-PN acts as the ENDS initiation rate from never users and ENDS-PF acts as the switching rate from current smokers.

Mexico’s gender-specific ENDS-PN and ENDS-PF by age group (ages 15-24, 25-39 and 40-65 years) were obtained from the 2015 Global Adult Tobacco Survey (GATS), the 2016 National Survey of Drugs, Alcohol, and Tobacco Consumption (ENCODAT) and the 2018, 2020, 2021 and 2022 National Health and Nutrition Survey (ENSANUT). ENDS-PN was calculated as those who answered “not at all” to “In the past, have you smoked tobacco on a daily basis, less than daily or not at all?” and answered “daily or less than daily” to “Do you currently use electronic cigarettes on a daily basis, less than daily, or not at all?” divided by the total number of never smokers. ENDS-PF was calculated as those who answered “Not at all” to “Do you currently smoke tobacco on a daily basis, less than daily, or not at all?”; “daily or less than daily” to “In the past, have you smoked tobacco on a daily basis, less than daily or not at all?” and answered “daily or less than daily” to “Do you currently use electronic cigarettes on a daily basis, less than daily, or not at all?” divided by the total number of former smokers. To fit the model, we transformed survey data by age group (15-24, 25-39 and 40-65 years) into single-age ENDS-PN and PFs by applying the age group’s value to each age within each group. We also estimated ENDS-PN and ENDS-PF values for years without survey information (2017 and 2019) by averaging the proportions from neighboring years (i.e., we used 2016 and 2018 to estimate 2017 and 2018 and 2020 for 2019). Mexico SAVM assumes that ENDS-PN and ENDS-PF remain constant at the 2022 level (last year of survey data) from 2023 to 2049, effectively assuming that ENDS initiation and cessation offset each other. We decided on this assumption given the limited information on ENDS initiation and cessation patterns under the current context in Mexico where products are banned yet available and to keep the model manageable with the least number of assumptions. S1 Table presents the ENDS-PN and ENDS-PF values from surveys used as inputs.

To derive Mexico-specific mortality rates by smoking status (never, current and former), SAVM uses the product of the ratio (R) (R= overall Mexico mortality rates/overall US mortality rates) and US death rates (e.g., *Mexico never death rates = R * US never death rates*). The US male (female) overall mortality rate was 0.12% (0.04%) at age 20, increasing to 0.21% (0.14%) at age 40 and 1.13% (0.68%) at age 60 in 2009. It decreases in relative terms by 46% (43%) from 2009-2049 for males (females) at age 20, 52% (49%) for age 40, and 48% (47%) for age 60. For smokers who quit before age 35, SAVM assumes the same mortality risks as never smokers (50–52). US smoking status and mortality data are obtained from the CISNET Lung group (50, 53–55). The CISNET mortality rates are based on mortality data through 2012 and smoking relative risks informed by data through the early 2000s, prior to the wide adoption of ENDS (56).

### Mexico SAVM calibration and validation

We first calibrated the ENDS-Restricted Scenario to ensure that projected future trends in Mexico were consistent with observed Mexican current smoking trends. Projected changes in smoking prevalence (smoking relative reductions) between 2009 and 2015 are compared to those changes from the Mexican GATS 2009 to 2015 for all population (15 to 65 years old) and three age groups (15 to 24, 25 to 39 and 40 to 65). We utilized GATS for Mexico SAVM calibration because it predates the widespread adoption of ENDS in Mexico, offering a clearer picture of pre-e-cigarette smoking prevalence. Based on the discrepancy between the model and the surveys in 2009-2015, we calibrated the model by applying separate smoking initiation and smoking cessation adjusters, i.e., a fixed scaling factor (adjuster) to both the ENDS-Restricted Scenario smoking initiation and cessation rates. The Mexico SAVM smoking initiation adjusters are set for males (females) at 1.52 (1.53) for ages 15-24 years and at 1.78 (1.56) for ages 25-40 years of the smoking initiation from the ENDS-Restricted Scenario. Smoking cessation adjusters are set at 0.50 (0.50) for ages 25-39 years and 0.48 (0.00) for ages 40 years and above of the smoking cessation from the ENDS-Restricted Scenario.

Mexico SAVM initially underestimated exclusive ENDS prevalence (from never and former smokers) use for ages 15 to 24 years for males and females. Calibration to ENDS estimates for this age group was made by replacing the original ENDS-PN or ENDS-PF from the Mexican surveys with the product of [*Exclusive ENDS prevalence from surveys for each data point/Prevalence estimates of Exclusive ENDS use from SAVM without calibration] multiplied by the corresponding ENDS-PN or ENDS-PF from each year*]. See S1 Table for the adjusted ENDS-PN and ENDS-PF.

The validation data used for the Mexico SAVM was over a specified period based on survey availability. The Mexico SAVM was validated by comparing relative changes in current smoking prevalence between 2015 and 2018 and 2015 and 2022 from the ENDS-Restricted Scenario to those from ENSANUT 2018 and 2022. These surveys were chosen because they collect nationally representative data representing tobacco and ENDs use trends in Mexico under the current national ENDS ban. They enabled us to estimate current smokers using the same measure as from GATS 2009 (baseline inputted data).

### The ENDS-Unrestricted Scenario

The ENDS-Unrestricted Scenario projects the prevalence of never, current, and former smokers, current exclusive ENDS users, and former ENDS users while allowing for direct switching from smoking to exclusive ENDS use (Fig 2), assuming that the current ENDS-ban is lifted in 2024. The baseline transition rates (switching rates) from smoking to exclusive ENDS use before age 35 and to former smokers using ENDS after age 35 are based on US cigarette and ENDS regular switching rates through 2017 (57), implying a relatively unrestricted approach to ENDS, with strong tobacco control policies. The unrestricted case implicitly assumes that ENDS regulations are implemented at a national level and strongly enforced, similar to enforcement in the UK and Canada (58). Later the transition rates and assumptions are varied in sensitivity analyses.

**Fig 2.**
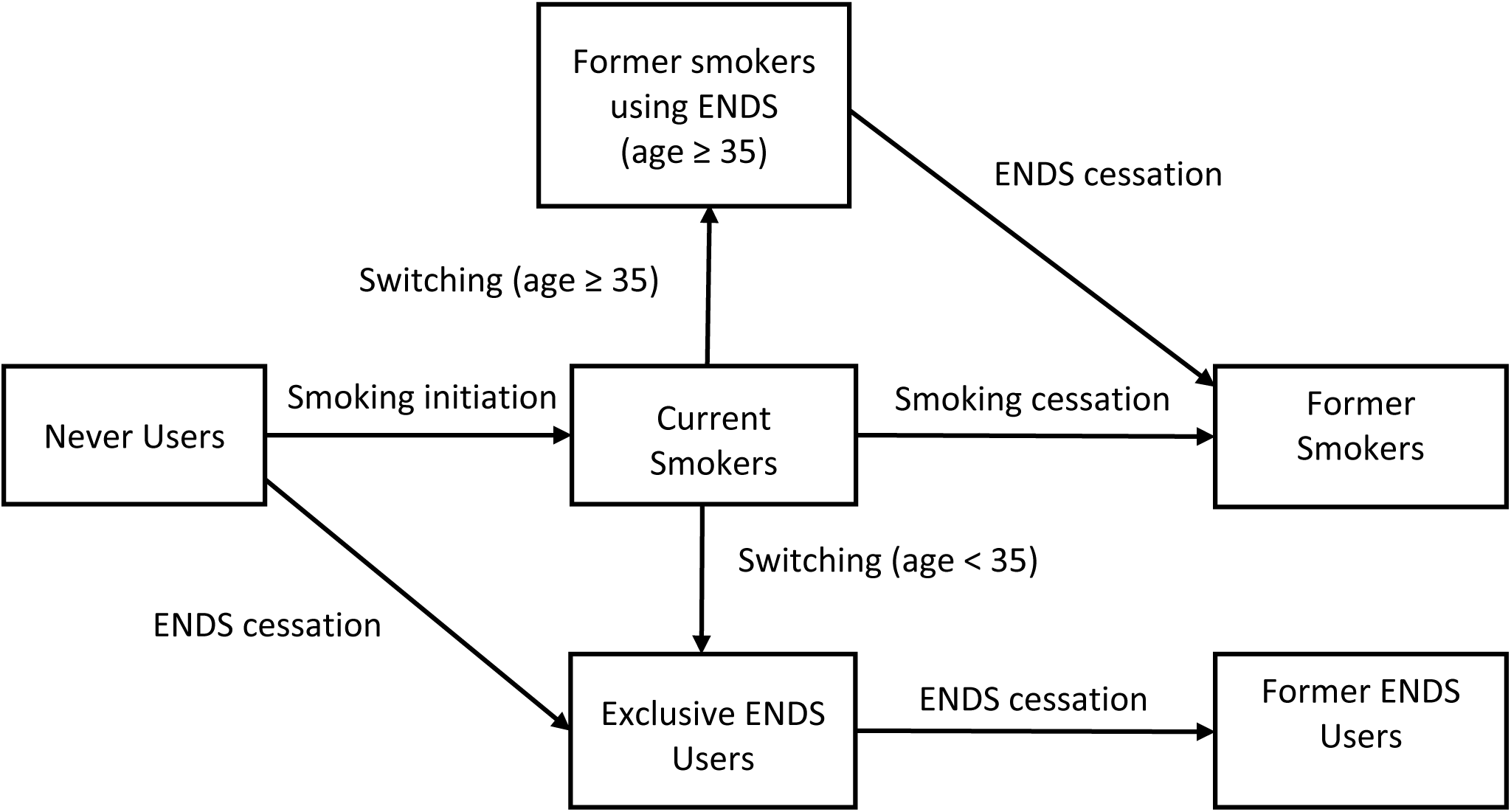
Transitions between Smoking and ENDS Use in the ENDS-Unrestricted Scenario

The ENDS-Unrestricted Scenario smoking and ENDS initiation rates were developed by applying separate smoking and ENDS use multipliers to the ENDS-Restricted Scenario smoking initiation rates, thus implying an age and gender pattern for initiation like those in the ENDS-Restricted Scenario subject to the constant scaler (multiplier). A smoking initiation multiplier >100% implies that smoking initiation with ENDS legalized availability increases above that in the presence of an ENDS ban (i.e., a gateway into smoking). Less than 100% implies ENDS diverting the never smokers away from smoking initiation. The Mexico SAVM smoking initiation multiplier is set at 75% of the smoking initiation from the ENDS-Restricted Scenario. The ENDS initiation rate multiplier is 50% of the ENDS Restricted Scenario smoking initiation. These parameters are based on the rapid decline in US youth and young adult smoking since vaping increased (59–61) and consider Mexico vaping trends. Mexico SAVM assumes that the ENDS-Unrestricted Scenario initiation rate multipliers for both genders and all ages are constant over time.

As mentioned, the ENDS-Unrestricted Scenario allows individuals to quit smoking and switch to exclusive ENDS use (aka switching rate). Prevalence of ENDS use is higher in the US than in Mexico (30, 31), consequently the US population has a higher exchangeability between ENDS and cigarette use (62, 63). Further, ENDS regulation in the US is less restrictive than in Mexico (14). To account for these differences, Mexico SAVM switching rates are set as 50% of the rates from prospective data from the US PATH survey over 2013-2017. We estimate male (female) switching rates as 2.0% (1.3%) per year for ages under 24, 1.3% (1.0%) for ages 25-34, 1.3% (0.8%) for ages 35-44, 0.7% (0.7%) for ages 45-54, 0.6% (0.7%) for ages 55-64 and 0.3% (0.5%) for ages 65. These rates are applied each year. We conduct a sensitivity analysis and consider switching rates at 25% from US PATH rates to reflect the uncertainty on switching from cigarettes to ENDS when permissive cigarette regulations are in place (e.g., legalization of flavored tobacco-products) (64, 65). We also consider 75% of those switching rates from PATH to showcase a scenario where strong cigarette policies and strong enforcement are in place.

Smoking cessation multipliers reflect those who quit both smoking and ENDS use. With ENDS unrestricted availability, smoking cessation rates of those who do not continue to vape regularly or who quit smoking without vaping are maintained at 100% of the ENDS Restricted smoking cessation rates. In contrast to the Restricted Scenario, the Exclusive ENDS and Former smokers-ENDS users can quit ENDS use through ENDS cessation rates and become former ENDS users. We assume the ENDS cessation as 100% of the ENDS Restricted smoking cessation rates.

An ENDS relative risk multiplier specifies the ENDS excess mortality risks relative to the excess mortality risk for current and former smokers (*current or former smoker death rate – never smoker death rate*). We consider a constant ENDS excess mortality risk at 15% that of smoking for both genders at all ages, based on estimates reached through a multi-criteria decision analysis (66) and an independent review (67). In Mexico SAVM, people who quit smoking but currently vape are accorded the risk of former smoking plus the ENDS risk multiplied by the difference in risk between current and former smokers. To reflect uncertainty around this estimate (68–70), we performed a sensitivity analysis by also considering ENDS relative risks of 5% and 25% of excess risks of smoking.

### Public Health Impacts

The public health impact of the ENDS-Unrestricted Scenario is evaluated as the difference in projected smoking- and vaping-attributable deaths (SVADs) and life-years saved (averted life-years lost [LYLs]) between the Restricted and Unrestricted Scenarios for individuals aged 15-65 years. Based on previous approaches (54, 71), SVADs are calculated by multiplying the number of people who currently (formerly) smoked and vaped by their excess mortality rate, measured by the current (former) smoking and vaping minus never smoking mortality rates. LYLs are calculated by multiplying the number of SVADs by the remaining life years of a never-smoker at the same age.

## Results

### Validation of Current Smoking Prevalence in the Restricted ENDS Scenario

Compared to ENSANUT 2018 and 2022 surveys, for ages 15-65, Mexico SAVM under the Restricted Scenario projected that male (female) smoking prevalence remains stable from 25.6% (8.6%) in 2015 to 25.7% (8.8%) in 2022, while Mexican observed data reported an increase from 26.2% (8.8%) in 2015 to 29.1% (10.5%) in 2022. For males ages 15-24, Mexico SAVM predicts a slight increase in smoking prevalence with relative differences between years ranging from 2.4% by 2018 and 3.8% by 2022 contrasting with the reported decrease in prevalence from Mexican data, with differences ranging from −10.4% to −16.8%. For ages 25-39, both SAVM and survey data showed increasing trends but reported differences in trend changes. Mexico SAVM reported relative differences ranging from 2.2% to 1.8% compared to differences of 17.6% by 2018 and 23.5% by 2022 from surveys. Male estimates for ages 40 to 65 from SAVM projected a constant decrease in smoking prevalence, but this pattern is not observed in the trend from surveys (Fig 3a).

**Fig 3a.**
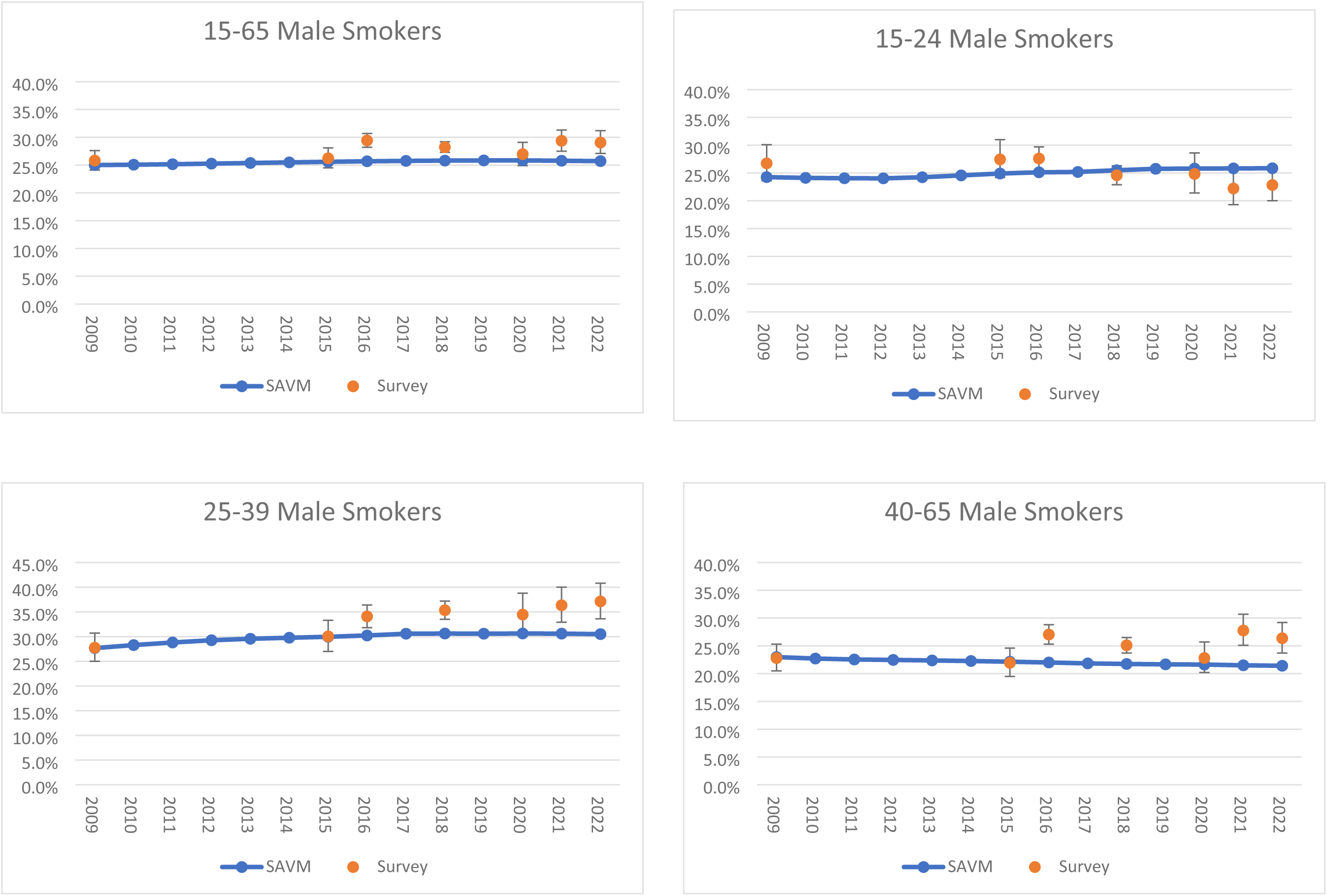
Validation of Mexico SAVM male current smoker estimates against Mexican surveys between 2015 and 2022.

For females, overall, SAVM under the Restricted Scenario underestimated smoking prevalence from surveys except for ages 45-65 years. For ages 15 −24 and 40-65, both Mexico SAVM and survey data smoking prevalence remained relatively stable after 2015, with SAVM projecting a slight increase in trends. The largest relative differences between SAVM and the surveys were observed for ages 25-39 years, with relative differences ranging from 5.7% to 3.4% from SAVM compared to differences of 22.5% by 2018 and 50.4% by 2022 from Mexico survey data (Fig 3b). Complete results by age group can be seen in S2 Table.

**Fig 3b.**
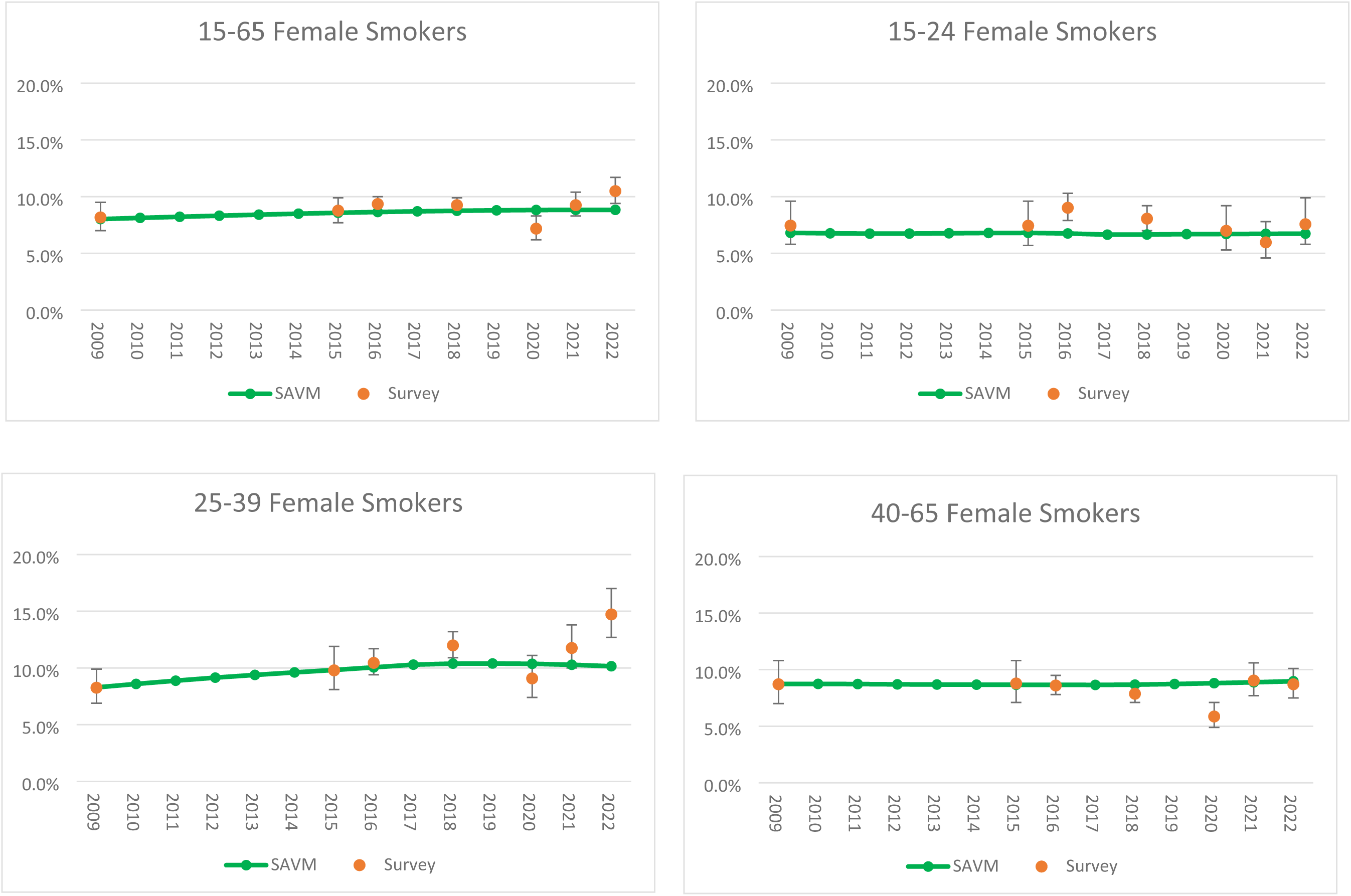
Validation of Mexico SAVM female current smoker estimates against Mexican surveys between 2015 and 2022.

### Comparison of ENDS-Restricted Scenario and ENDS-Unrestricted Scenario for All Cohorts

Table 2 presents the results by gender for all cohorts 15 to 65 years born in or after 2009 with an ENDS risk at 15% of excess smoking mortality risk.

**Table 2.** The Mexico Smoking and Vaping Model, ENDS Restricted vs. Unrestricted Scenario by gender with a 15% ENDS Risk Multiplier, All Cohorts with New Births, Ages 15-65, 2009–2049.

|  | Years | 2009 | 2015 | 2025 | 2035 | 2049 | Total |
| --- | --- | --- | --- | --- | --- | --- | --- |
| **Male** |  |  |  |  |  |  |  |
| **ENDS-Restricted Scenario** | **Smokers (%)** | 25.01% | 25.60% | 25.58% | 24.53% | 21.75% |  |
|  | **N-ENDS users (%)** |  | 0.36% | 1.07% | 0.99% | 0.91% |  |
|  | **FS-ENDS users (%)** |  | 0.04% | 0.11% | 0.11% | 0.12% |  |
|  | **SVADs** | 26,340 | 30,810 | 37,943 | 39,809 | 35,852 | 1,543,675 |
|  | **LYLs** | 655,037 | 780,571 | 962,664 | 1,008,628 | 886,537 | 38,981,126 |
| **ENDS-Unrestricted Scenario** | **Smokers (%)** | 25.01% | 25.60% | 24.98% | 19.03% | 13.03% |  |
|  | **N-ENDS users (%)** |  | 0.36% | 1.74% | 7.06% | 10.67% |  |
|  | **FS-ENDS users (%)** |  | 0.04% | 0.21% | 1.04% | 1.41% |  |
|  | **SVADs** | 26,340 | 30,810 | 37,750 | 36,938 | 28,422 | 1,452,634 |
|  | **LYLs** | 655,037 | 780,571 | 957,189 | 926,601 | 679,616 | 36,431,244 |
| **Health impacts** | **Smokers averted (%)** |  |  | -2.3% | -22.4% | -40.1% |  |
|  | **Averted SVADs** | - | - | 193 | 2,872 | 7,430 | 91,041 |
|  | **Life-year gains** | - | - | 5,475 | 82,027 | 206,920 | 2,549,883 |
| **Female** |  |  |  |  |  |  |  |
| **ENDS-Restricted Scenario** | **Smokers (%)** | 8.02% | 8.58% | 8.84% | 8.73% | 8.53% |  |
|  | **N-ENDS users (%)** |  | 0.12% | 0.67% | 0.62% | 0.59% |  |
|  | **FS-ENDS users (%)** |  | 0.02% | 0.11% | 0.10% | 0.10% |  |
|  | **SVADs** | 3,374 | 5,106 | 7,083 | 6,876 | 8,043 | 283,539 |
|  | **LYLs** | 86,046 | 127,677 | 167,704 | 167,694 | 186,037 | 6,818,611 |
| **ENDS-Unrestricted Scenario** | **Smokers (%)** | 8.02% | 8.58% | 8.67% | 7.22% | 5.89% |  |
|  | **N-ENDS users (%)** |  | 0.12% | 0.87% | 2.29% | 3.52% |  |
|  | **FS-ENDS users (%)** |  | 0.02% | 0.14% | 0.42% | 0.67% |  |
|  | **SVADs** | 3,374 | 5,106 | 7,044 | 6,446 | 6,683 | 268,058 |
|  | **LYLs** | 86,046 | 127,677 | 166,758 | 156,921 | 153,174 | 6,439,220 |
| **Health impacts** | **Smokers averted (%)** |  |  | -1.9% | -17.2% | -30.9% |  |
|  | **Averted SVADs** | - | - | 40 | 430 | 1,360 | 15,481 |
|  | **Life-year gains** | - | - | 946 | 10,773 | 32,863 | 379,391 |
| **Both genders** |  |  |  |  |  |  |  |
| **Health impacts** | **SVADS averted** |  |  | 233 | 3,301 | 8,790 | 106,522 |
|  | **LYLs averted** |  |  | 6,421 | 92,800 | 239,784 | 2,929,274 |
|  | **SVADS averted (%)** | - | - | 0.6% | 7.4% | 18.7% | 5.8% |
|  | **LYLs averted (%)** | - | - | 0.6% | 7.9% | 22.4% | 6.4% |

Abbreviations: ENDS =electronic nicotine delivery systems, LYL = Life years lost, SVADs = smoking and vaping attributable deaths.
<sup>a</sup> Cumulative results include the deaths and life-years lost, which are the sum of attributable deaths or life-years lost over the years 2009–2049.
<sup>b</sup> Restricted Scenario refers to values in the assumption of restricted ENDS use.
<sup>c</sup> Unrestricted Scenario refers to values relative to less restricted ENDS use.
<sup>d</sup> Difference between the ENDS Restricted Scenario and ENDS Unrestricted Scenario includes the percent averted smokers (measured by the relative reduction in the smoking prevalence each year) by gender, averted SVADs, and LYLs for males, females, and both genders. The relative differences (%) of averted SVADs and LYLs are also available for both genders by using formulas:
SVADs averted (%) = $\text{SVAD averted} / \text{SVAD}_{\text{Restricted ENDS}}$ ; LYLs averted (%) = $\text{LYL averted} / \text{LYL}_{\text{Restricted ENDS}}$

In the ENDS-Restricted Scenario, adult male smoking prevalence is projected to decline from 25.6% in 2015 to 24.5% in 2035 and 21.8% in 2049. Female smoking prevalence remained stable from 8.0% in 2015 to 8.7% in 2035 and 8.53% in 2049. Male and female exclusive ENDS use was projected an increase for males (females) from 0.36% (0.12%) in 2015, 1.07% (0.62%) in 2035 and 0.91% (0.59%) in 2049. From 2009-2049, SAVM projects for males (females) 1.5 (0.28) million cumulative SVADs with 39.0 (6.8) million cumulative LYLs due to smoking and vaping.

In the ENDS-Unrestricted Scenario, by 2035 smoking prevalence declines to 19.0% for males and 7.2% for females. These values represent a relative decline of 22.4% in male and 17.2% in female smoking prevalence from the values under the ENDS-Restricted Scenario. By 2049, male smoking declines to 13.0%, a 40.1% relative decline while female smoking declines to 5.9%, a 30.9% relative decline from the Restricted Scenario. Under a ban removal, exclusive ENDS use projected an increase for males (females) to 7.1% (2.3%) in 2035, and to 10.7% (3.5%) in 2049. Mexico SAVM projected smoking and vaping prevalence by age-group and gender in two scenarios are shown in S3 Table. The prevalence of former smokers using ENDS is projected to increase to 1.4% for males and 0.7% for females by 2049. From 2009-2049, a total of 1.5 million male and 0.27 million female SVADs and 36.4 male and 6.4 million female LYLs are projected.

From 2025-2049, approximately 106.5 (91.0 males and 15.5 females) thousand SVADs are averted, and 2.9 (2.5 males and 0.4 females) million life years are gained in the ENDS-Unrestricted Scenario compared to the ENDS-Restricted Scenario, representing relative reductions of 5.8% in SVADs and 6.4% in LYLs.

### Sensitivity Analysis

Sensitivity analyses are shown in S4 and S5 Tables for SVADs and LYLs for both genders combined for the ENDS-Restricted and Unrestricted Scenarios for 2025-2049. To gauge the public health effect of the change in parameters, we estimate relative changes from the baseline scenario (i.e., ENDS relative risk of 15% that of cigarettes). Applying the same input parameters but with 25% of the US PATH switching rates, public health gains decline by −43.9% to 59,748 SVADs averted and by −43.1 % to 1.7 million LYLs averted compared to 106,522 SVADS and 2.9 million LYLs averted with a 50% from US PATH switching rates. In contrast a 75% of the US PATH switching rates estimated 148,695 SVADS and 4.1 million LYLs averted, representing an increase in almost 40% of the health impact from baseline. In a separate sensitivity analysis, mortality relative risks associated with ENDS were examined, ranging from 5% to 25% of that for cigarettes. The total number of averted SVADs for both genders decreased from 121,375 at the 5% ENDS risk level to 106,522 at 15% and further to 92,806 at 25% risk, indicating a relative reduction of 24.3% from the lower bound (5%) to the upper bound (25%). Total averted LYLs decline from 3.5 million with 5% ENDS risk to 2.9 million with 15% risk and 2.5 million with 25% risk, a 24.9% relative reduction from the lower to the upper bound.

## Discussion

Our simulation study indicates that lifting the ENDS ban in the upper middle-income country of Mexico could reduce cigarette use and its negative public health impacts. Results suggest that implementing less restrictive ENDS regulations could decrease smoking prevalence by 40.1% in males and 30.9% in females by 2049 compared to continuing a national ENDS ban. This reduction in prevalence would save 2.9 million life-years and avert almost 106,000 deaths between 2025 and 2049. These results align with those from simulation models on ENDS ban removal from HICs, whether estimated using previous SAVM (i.e., The Australia SAVM) (72) or alternative approaches (22). Our results are also consistent with a systematic review of modeling studies on public health impacts of e-cigarettes in HICs (20), which concluded that a population increase in ENDS use due to increased ENDS availability would be associated with decreased smoking rates, smoking-associated mortality, and health system costs, as well as increased QALYs compared to a scenario where combustible cigarette was the only product in the market.

Previous simulation models for Mexico (SimSmoke) focus on the public health impacts of tobacco control policies focusing on cigarettes. For example, the model estimated that a one-peso increase in the cigarette tax could avert 146,000 female deaths and 483,000 male deaths, along with gains of 2.9 million and 9 million life-years, respectively (73). Mexico SimSmoke also estimated that stricter enforcement of existing policies could lead to relative reductions in daily smoking prevalence by 25.6% for men and 26.7% for women, with nondaily smoking also declining by 26.9% and 27.3%, respectively (74). However, while Mexico SimSmoke effectively captured the decline in daily smoking following the implementation of various FCTC policies, it failed to anticipate the concurrent increase in nondaily smoking. This discrepancy highlights the necessity for further research to unravel the intricacies of nondaily smoking patterns and their implications for our current study. While this issue presents a challenge, it is crucial to acknowledge that lifting the ban on ENDS would complement the impact from other public health measures, and according to Mexico-SAVM, could save 106,000 lives and gain 2.9 million life-years over 24 years. This potential benefit should not be overlooked, even if it is somewhat attenuated by persistent nondaily smoking. Research is needed to understand whether switching to ENDS is more or less likely among Mexicans who smoke with varying frequency.

Despite Mexico’s de facto 2008 ENDS ban, national health surveys have reported observable utilization of e-cigarettes since 2015. Simultaneously, the prevalence of last 30-day smoking has remained stable (4, 31). Could lifting the ban reduce smoking prevalence? Current access to e-cigarettes in Mexico is influenced by two factors: low enforcement of the ENDS ban (14, 33) and a large informal economy (approximately 29% of GDP)(75) that appear to bolster their availability and affordability (4, 76, 77). Therefore, lifting the ban could have a relatively limited impact on overall cigarette use if it does not further increase access to and affordability of ENDS. Nevertheless, by legalizing and regulating ENDS, Mexican smokers could have access to more standardized and, likely, less harmful ENDS than those currently on the market, which are often mislabeled and include a range of potentially harmful constituents (78, 79). Moreover, current smokers might be more willing to try ENDS for cessation if these are approved and regulated than in the current environment.

Nonetheless, permissive ENDS regulation also carries risks, such as the potential renormalization of nicotine use attracting new users, particularly youth. To help prevent these downsides, for example, regulations could limit ENDS flavors that appeal to youth and young adults. To optimize the potential impact of legalizing ENDS, their regulation should be viewed as a component of a comprehensive strategy alongside strong tobacco control policies and enforcement to steer consumers away from cigarette smoking. Specifically, regulations should ban flavor capsule cigarettes, which come in a range of flavors as ENDS and are particularly popular among youth, women, and nondaily smokers (64), with increasing use rates (65, 80). It is also imperative for Mexico to ratify and enforce the Protocol to Eliminate Illicit Tobacco Products (71, 72], as its enforcement would play a pivotal role in curbing access to and availability of tobacco and nicotine products that pose potential harm to public health. Despite such efforts, it is unclear how the illegal market would respond to ENDS legalization, though it is unlikely that it would disappear altogether, given the size of the informal economy in Mexico.

Research is needed to better project such market dynamics. Ultimately, the decision to lift the ban hinges on a balancing act. Policymakers must carefully weigh the potential health gains for existing smokers against the risks of increased nicotine use, especially among young people. Implementing stringent regulations, including marketing restrictions, age verification, and flavor bans, could mitigate these risks. Additionally, ongoing research and monitoring are crucial to understanding the long-term impact of lifting the ban on both individual and public health in Mexico.

### Limitations

SAVM’s accuracy depends on the availability of tobacco surveillance data to exhibit changes in smoking behaviors. Nationally representative surveys on e-cigarette use in Mexico started in 2015; this short period of data collection, in combination with the COVID-19 pandemic in the last years, brings uncertainty to the stability of our estimates. As more recent data are published and Mexico SAVM is updated, the projections in this paper should be evaluated and updated if necessary.

Further consideration should also be given to the assumptions of the ENDS-Restricted and Unrestricted Scenarios. The ENDS-Restricted Scenario applies initiation and cessation rate parameters based on an age-period-cohort smoking analysis (49, 81, 82) using data from Mexico through 2016 and did not consider changes in use since 2016. Further, the data on smoking prevalence were based on any cigarette use and thus may overestimate the effect on established cigarette use (i.e., smoked at least 100 cigarettes during their lifetime and currently smokes daily or nondaily), the more appropriate population in gauging public health impacts (57, 83). We also calibrated the model to Mexican trends between 2009 and 2015, dampening recent increases in ENDS use among 15–24-year-old Mexicans and flattening their declining smoking rates. These modifications reduced the decline in smoking rates and factored a smaller increase in e-cigarette use for individuals under 40 years than observed in recent years. In addition, Mexico SAVM projected that up to 2049, ENDS use among former smokers remains under 2%. SAVM classifies individuals who quit smoking before age 35 as never smoking due to the reduced mortality risk; this could underestimate the actual prevalence of ENDS use among former smokers. Finally, we do not report results on cigarette users aged 66 and above despite their minimal vaping prevalence. We expect ENDS’s impact on this older population will occur in those who become older after 2050; still, this demographic presents an opportunity for the potential adoption of ENDS (84).

Our analysis of the ENDS-Restricted Scenario also assumed that the 2022 rates of exclusive ENDS use would continue into the short term. Recent evidence indicates that ENDS use in Mexico has generally continued to increase (30, 31). To the extent that ENDS use, especially exclusive use trends continue, the impact of eliminating Mexico’s ENDS ban is likely to be less than projected by our estimates because those likely to use ENDS may have already become regular users. Currently, ENDS use occurs predominantly among individuals who concomitantly use cigarettes (80% of males and 90% of females who use ENDS in 2021) (85), while the prevalence among never tobacco users is very low (31), indicating that the ban may have discouraged smokers from switching entirely to ENDS use, potentially due, at least partially, to the lack of legally available products and current cigarette policies.

SAVM follows a conservative approach and does not distinguish dual use from exclusive smoking and assumes the same cessation and switching patterns and health risks among dual users as among exclusive smoking (86, 87). However, this approach poses the risk of oversimplifying the complex behavior among individuals who use cigarettes and vape. While evidence of the health impact of dual relative to exclusive cigarette use is mixed (88–91), the health impacts of dual use merit further study. Further examination of dual use in Mexico and the impact of implemented tobacco regulations is warranted to inform more targeted and effective public health policies tailored to address the complexities of dual tobacco use behaviors. We also did not consider heated tobacco product, used by 1% of smokers according to a recent study (92).

The estimated number of averted deaths due to a more permissive ENDS policy is sensitive to assumptions about the presumed relative risks of ENDS use compared to smoking and the estimates of switching from smoking to ENDS (21). The ENDS-Unrestricted Scenario switching rates draw on US data from before 2017, a period when e-cigarette use had not yet reached its peak, and newer devices like nicotine-salt pods and disposable vapes (e.g., Puffbar) had not yet been released (57). Our projections scaled down to 50% tof he US switching rates to partially reflect the differences in ENDS consumption and tobacco control policies implementation and enforcement between the US and Mexico It also assumes a more stringent cigarette-oriented policy framework than what is currently implemented in Mexico. Presently, Mexico’s tobacco regulations afford consumers access to various combustible alternatives aside from ENDS, such as flavored capsule products, thereby offering a selection beyond vaping alone (33, 93). In the Unrestricted-ENDS Scenario, this expanded selection of combustible options would likely lessen the relative appeal of ENDS, potentially impeding the transition from smoking to vaping and decreasing the switching rates lower than 50%. This shift would consequently lead to a reduction in health gains by 43%, as estimated by our sensitivity analysis utilizing 25% of US PATH switching rates (S4 Table). Still, a deeper understanding of e-cigarette switching behavior is crucial, as it has the most significant impact on our public health projections (57, 72, 94). Moreover, attributing a conservative 15% ENDS excess risk highlights the potential for even greater health risks from e-cigarettes regardless of the ban status. We performed a sensitivity analysis with the ENDS excess risk at 5% and 25% and the results showed that even if the risk from ENDS is higher than assumed there could be an important public health benefit of legalization.

In modeling the impact of a more permissive ENDS policy, we did not consider differences in past ENDS-oriented and cigarette-oriented policies in Mexico compared to the US. Negative government and health organization messaging and history of restrictive ENDS policies may reduce the impact of more permissive ENDS policies (95, 96). For example, messaging about the unknown health risks of ENDS may have augmented misperceptions of their harm relative to cigarettes. However, Mexico’s current cigarette-oriented policies may instead heighten the impact of more relaxed ENDS policies. For example, higher levels of cigarette taxes in Mexico (68%) relative to the US (40%), a national smoke-free-air law that was recently strengthened, and more restrictive cigarette marketing regulations in Mexico (93) along with lower incomes (97), may more strongly motivate current and potential future to switch to ENDS in Mexico, particularly if their price compared to cigarettes is lower than in the US.

Mexico-SAVM represents, to our knowledge, the first simulation modeling study of the potential effect of a permissive ENDS regulation in LMICs. Further study of the potential impact of alternative nicotine delivery products in LMICs is needed since they represent about 80% of the world population and carry most of the smoking-associated disease burden (2). However, a general lack of data on ENDS use from LMICs represents a limitation to quantifying the effects in these countries (3).

## Conclusion

Results from the Mexico SAVM suggest that greater access to ENDS and a more permissive ENDS regulation could create significant reductions in smoking prevalence and replace smoking with vaping. The results are subject to the model’s assumptions and uncertainty about the impact of countries switching from a more to a less restrictive ENDS regime and the role and level of cigarette-oriented policies. Nevertheless, the use of ENDS in low- and middle-income nations merits closer scrutiny, and further consideration should be given to the role of ENDS restrictions.

## Data Availability

Data used to parameterize the model are publicly available https://ensanut.insp.mx/ https://www.gob.mx/conapo/documentos/catalogo-digital-direccion-de-analisis-estadistico-e-informatica?idiom=es The SAVM package and User Guide are made available to the public by the University of Michigan and Georgetown University-Tobacco Center of Regulatory Science (TCORS)-Center for the Assessment of Tobacco Regulation (CAsToR) group upon request at: https://tcors.umich.edu/Resources_Download.php?FileType=SAV_Model

https://ensanut.insp.mx/

https://tcors.umich.edu/Resources_Download.php?FileType=SAV_Model

## Authors Contributions

Conceptualization: James F. Thrasher, Luz María Sánchez-Romero, Rafael Meza, David T. Levy

Formal analysis: Luz María Sánchez-Romero, Yameng Li, Luis Zavala-Arciniega

Funding acquisition: James F. Thrasher

Investigation: Luz María Sánchez-Romero, Luis Zavala-Arciniega, Katia Gallegos-Carrillo

Methodology: Luz María Sánchez-Romero, Yameng Li, David T. Levy

Resources: Luis Zavala-Arciniega, James F. Thrasher, David T. Levy

Writing-original draft: Luz María Sánchez-Romero, Yameng Li, David T. Levy

Writing-review & editing: Luis Zavala-Arciniega, Katia Gallegos-Carrillo, James F. Thrasher, Rafael Meza.

### Ethics approval and consent to participate

This study uses publicly available data sets. No ethical approval was required. This study uses publicly available anonymized data sets.

Informed consent was not required.

### Consent for publication

Not Applicable

### Availability of data and materials

Data used to parameterize the model are publicly available https://ensanut.insp.mx/https://www.gob.mx/conapo/documentos/catalogo-digital-direccion-de-analisis-estadistico-e-informatica?idiom=es

The SAVM package and User Guide are made available to the public by the University of Michigan and Georgetown University-Tobacco Center of Regulatory Science (TCORS)-Center for the Assessment of Tobacco Regulation (CAsToR) group upon request at: https://tcors.umich.edu/Resources_Download.php?FileType=SAV_Model

### Funding

Research reported in this publication was partly supported by the Fogarty International Center of the National Institutes of Health under award number R01 TW010652. We also acknowledge support from NIH/NCI grants U01CA253858 and K01CA260378. The content is solely the responsibility of the authors and does not necessarily represent the official views of the National Institutes of Health.

